# Prevalence and risk factors of Schistosomiasis among school aged children in Al- Fashir, North Darfur state, Sudan: a cross sectional study

**DOI:** 10.1101/2022.04.10.22273476

**Authors:** Ishraga Adam Elzain, Hamid Suliman Abdalla, Abeer Babiker Idris, Nagla Mohamed Ahmed, Salah Jomaa, Mohamed A. Hassan

## Abstract

**Objective:** Schistosomiasis represents a significant health problem in Sudan. School aged children who live in areas with poor sanitation are often at risk because they tend to spend time swimming or bathing in water containing infectious cercariae. Therefore, this study aimed to investigate schistosomiasis in terms of prevalence of the infection, and its risk factors among school aged children at Al- Fashir, the capital city of North Darfur state in Sudan.

**Results:** In this study, *S. haematobium* was detected in 6.1% of the school age children at Al- Fashir. Also, hematuria was detected in 85.7% of infected patients, and there was significant correlation between hematuria and presence of *S. haematobium* eggs (P. value= 0.001). Regarding the risk factor, the low prevalence rate of *S. haematobium* was observed in populations who depend on faucets as water sources and live in Nifasha and Zamzam camps.

## 1. Introduction

*Schistosoma* is a genus of trematodes include parasitic flatworms which responsible for a highly significant group of infections in humans and animals termed schistosomiasis. Schistosomiasis is considered by the World Health Organization (WHO) as the second-most socioeconomically important parasitic disease after malaria partially in tropic area (1, 2). It is estimated that 250 million of people are infected and approximately 700 million are at risk of infection (3). It is widely acknowledged that schistosomiasis infection prevalence and intensity curves show peaks in children aged 6–15 years (4, 5). There are two forms of schistosomiasis which known as urinary schistosomiasis caused by *S. haematobium* and intestinal schistosomiasis caused by *S. mansion* and *S. japonicum*. However, snails act as intermediate hosts and require for the development of the cycle to produce the cercaria, the infective stage (6). The main approaches to control schistosomiasis include drug treatment of infected patients and snail control (6, 7).

There is no doubt that schistosomiasis infection has a global impact as a potential risk for human’s life, especially in tropical and semi tropical areas such as Sudan. In Sudan, *Schistosomiasis* remains a significant public health problem and is mostly occurring in Al-Gezera Agriculture Schemes (8). Studies on *Schistosoma* had started early in 1980 (9, 10). Although the available data shows high rate of *Schistosoma* transmission among the local Darfur population, studies that address *Schistosomiasis* infection in Darfur state are very few. However, to our knowledge, prevalence of schistosomiasis infection and its risk factors in Al-Fashir, the capital city of North Darfur state, has not been studied yet. Therefore, this study aimed to investigate schistosomiasis in terms of prevalence of the infection, and its risk factors among school aged children at Al-Fashir city. School aged children who live in areas with poor sanitation are most often at risk because they tend to spend time swimming or bathing in water containing infectious cercariae (11).

## 1. Materials and methods

### 1.1 Study design and study setting

This observational cross-sectional study was conducted throughout Al-Fashir, the capital city of North Darfur state in western Sudan. Samples were taken from different schools, health centers and camps. The collected residential areas sites include a) Tombase in the northern, b) Makkraka located in the southern, c) Awlad Elreef located in western of the city. While the camps Zamzam and Nifasha located northen and southern of the city, respectively. Water sources for daily activities include, tankers, wells, canals and tap water were fetched by donkeys and cars. There is a large pond in the center of Al-Fasher city called Foula, which is filled with rainwater every autumn season and gradually decreases and sometimes dries up in the winter season, and it contains *Bulinus* snails (Figur 1A). This canal is a source of water, especially for the people of the villages who come with donkeys to sell their simple trade in the market near the canal, and children swim in this water on their way back from school, especially in the summer, and the cars are washed on the sides of this canal.

**Figure 1.**
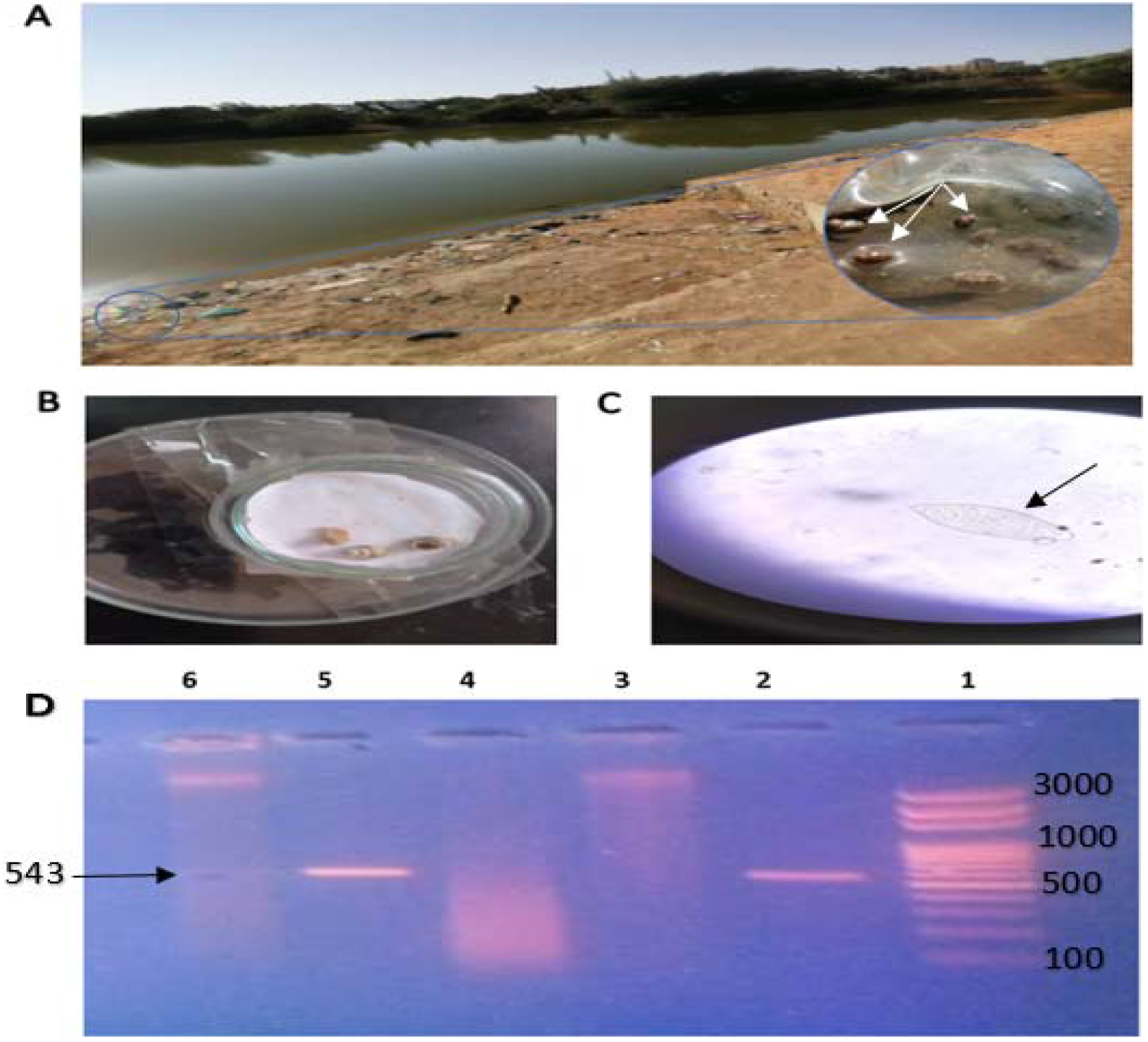
1A. The Foula pond in the central of Al- Fashir city. Arrows indicate *Bulinus* snails located at the edges of the Foula pond. 1B. Shows snails belong to *Bulinus* genus (the host of *S. haematobium*). 1C. Microscopical view of *S. haematobium*. 1D. 1% Agarose gel result of *COX1* gene amplification. Lane 1: 100 bp DNA ladder. Lane 2 and 5: positive result (543 bp). Lane 3 and 6: non-specific bands. Lane 4: negative result.

### 1.2 Sample Collection

All the study participants were requested to provide both urine and stool samples by using dry, clean well-labelled plastic containers. Approximately, 5–10 ml of urine and 5–7 g of feces was collected. The sample size was determined according to the following equation:

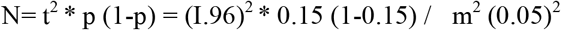

For the calculation, 95% confidence level (t), 15% prevalence of the disease (p) and 5% margin of error (m) were used. Therefore, the sample size (N), for urine and stool samples, was 195 for each. In addition, the sociodemographic and associated risk factors were obtained via a standardized questionnaire.

### 1.3 Macroscopic examination

Urine specimens were examined macroscopically for their appearance and color. Hematuria and chemical properties were examined using reagent strips. Stool specimens were macroscopically examined by naked eye for their consistency, color, and worms.

### 1.4 Microscopic examination

Urine samples were microscopically examined for the presence of *S. haematobium* eggs by using simple centrifugation/sedimentation technique (12). Microscopic diagnosis of *S. mansoni* infection in fecal samples was performed using formal ether concentration method (13). All microscopic slides containing the eggs of *S. haematobium* or *S. mansoni* were considered positive, whereas the absence of the eggs was recorded as negative.

### 1.5 Molecular detection of *S. haematobium*

DNA was extracted from *Schistosoma* ova using G-DEXTM IIb Genomic DNA extraction kit. Then the extracted DNA was amplified using specific-species primer for *cytochrome oxidase subunit 1* (*COX1*) of *S. haematobium*, as previously described (14).

### 1.6 Statistical analysis

Data obtained from this study were entered into Statistical Package for Social Sciences (SPSS) version 23 and analyzed using Chi square to determine the correlation between variables and risk factors. P-value less than 0.05 was considered significant.

## 2. Results

### 2.1 Examination of urinary and fecal samples for *Schistosoma* in school aged children

Microscopic examination of 196 urine samples obtained from school age children revealed that eggs were found in 12 (6.1%), and the remaining 186 (93%) samples were clear (Figure 1). According to the morphological appearance of the eggs, it becomes clear that the causative parasites was *S. haematobium*. In addition, microscopic examination of 196 fecal samples did not reveal any ova of *S. mansoni* which indicate negative intestinal *schistosomiasis*. Regarding the molecular detection of *S. haematobium*, among the 12 positive urine samples by concentration technique, it was found eight of them were positive by PCR (Figure 1D).

### 2.2 Frequency of hematuria in urine samples

In this study, the frequency of hematuria samples was 14 (7.14%). Out of them, 12 (85.71%) had *S. haematobium*, and 2 (14.29%) did not harbor *S. haematobium* eggs (negative). The remaining 182 urine samples, which have no hematuria, were also negative for the parasite. There was significant association between hematuria and infection of urinary *schistosomiasis*.

### 2.3 Study population characteristics

As shown in Table 2., the study population was categorized according to their water supplies into different sources which included tankers, donkers (deliver water with donkeys), canals, and faucets. In this study, the prevalence of *S. haematobium* was most common in those who deepened on donkers as water supply 2/18 (11%) followed by canals 4/50 (8.0%). Whereas there was no *Schistosoma* infection among those who deepened on faucets as water supply. Regarding the residents of the study population, the *Schistosoma* infection was most common among those who live in Tombasie (14%) followed by Makraka (13.6%). While those who live in Awlad Alreef and Camps had no infection.

**Table 1.**
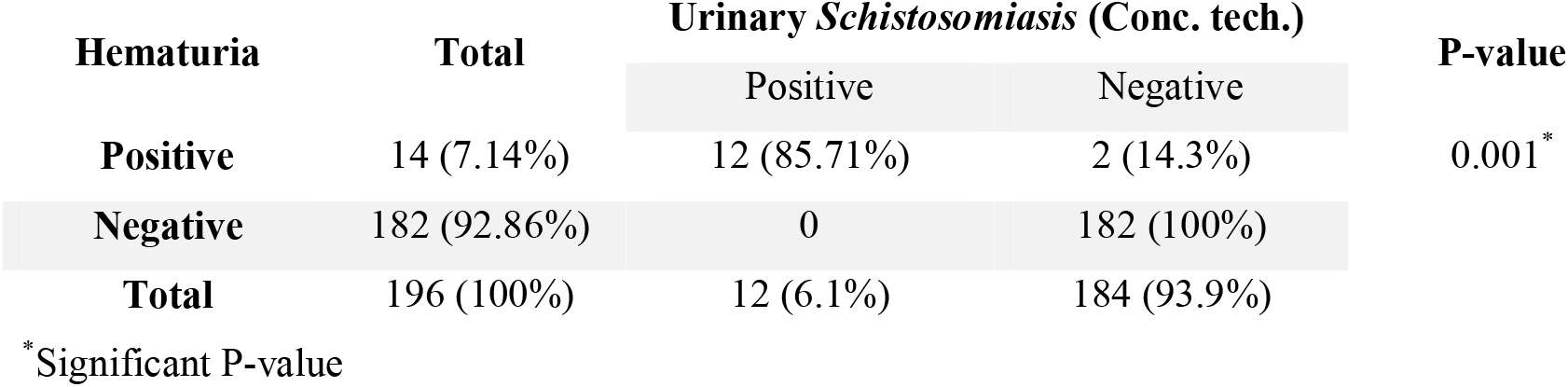
Association between hematuria and infection of Schistosoma

| Hematuria | Total | Urinary *Schistosomiasis* (Conc. tech.) |  | P-value |
| --- | --- | --- | --- | --- |
|  |  | Positive | Negative |  |
| Positive | 14 (7.14%) | 12 (85.71%) | 2 (14.3%) | 0.001* |
| Negative | 182 (92.86%) | 0 | 182 (100%) |  |
| Total | 196 (100%) | 12 (6.1%) | 184 (93.9%) |  |

\*Significant P-value

**Table 2.**
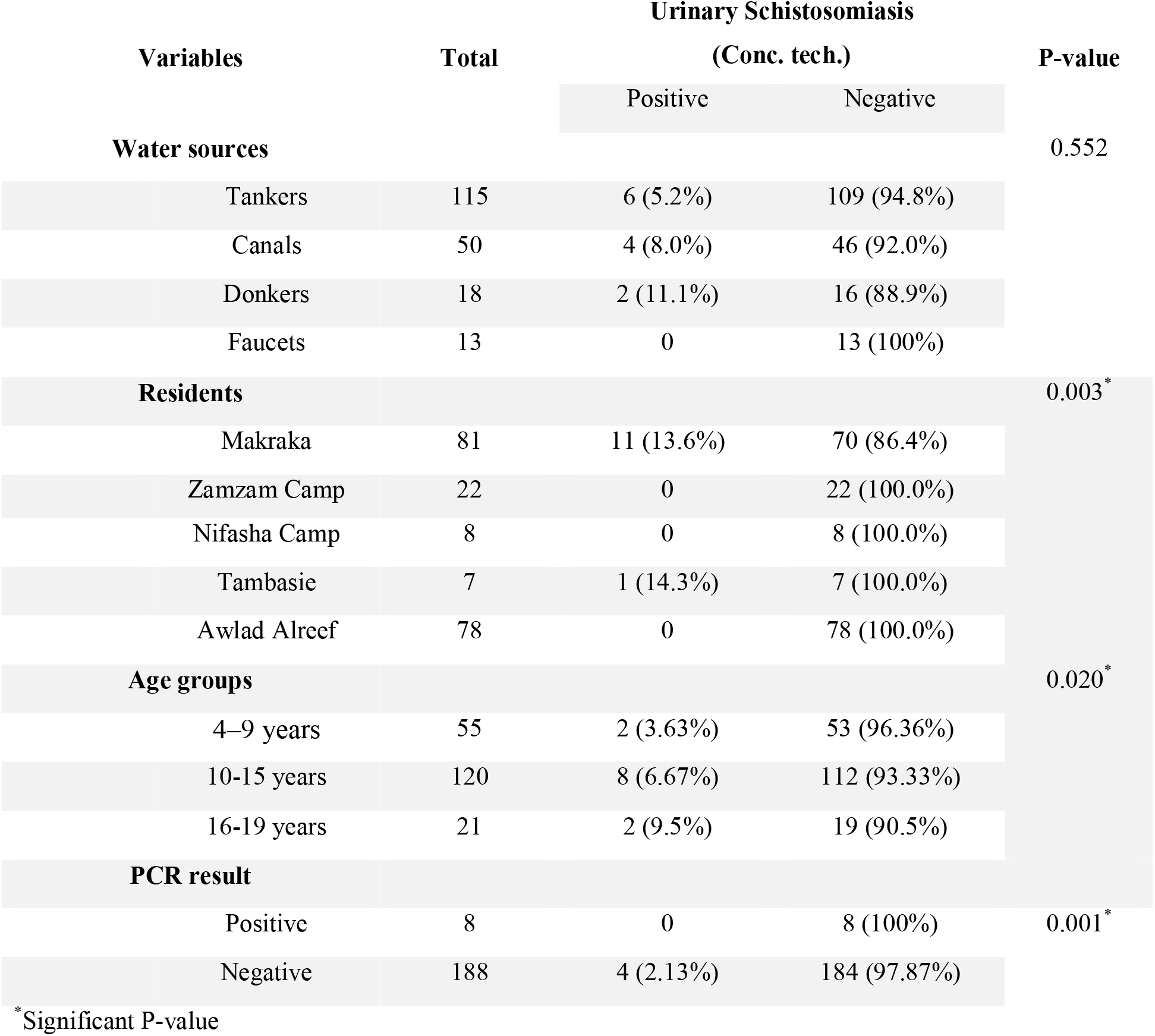
Association between frequency of urinary Schistosomiasis and population’s characteristics

In addition, the study population was grouped depending on their age as following: 4-9 years, 10-15 years and 16-19 years. However, the most common urinary schistosomiasis was found among the age group ranged from10 to 15 years old (66.7%) following by the age groups ranged from 16 to 19 years and 4 to 9 years old with 16.7 %, as represented in Table 2.

## 3. Discussion

In this study, *S. haematobium* was detected in 6.1% of the school age children at Al-Fashir. As there are no previous data conducted in this area, hence it could be regarded as a preliminary report on urinary schistosomiasis in Al-Fashir. Such absence of published studies made it difficult to compare the data with previous one. The study finding agrees with a study conducted in Egypt which found the prevalence of *S. haematobium* was 7.8% (15). However, it disagrees with studies conducted on various part of Sudan. It was higher than previous study performed in Al-Gezera areas which found the prevalence of *S. haematobium* was 0.9% (8). In contrast, the prevalence of *S. haematobium* in Al-Fashir was lower than in Um-Asher area, Southern Kordofan state, South Darfur (Alsafia and Abuselala cities) and White Nile River basin which were 12.9%, 23.7%, 56% and 45.0%, respectively (16-19). Such differences in the prevalence rate could be attributed to a numerous factor such as the variations in water sources, personal hygiene, and sanitation and environmental factors that affect the presence and distribution of snails (parasite’s host). Moreover, in the present study, mucus was found to occur in most of the stool samples. However, no presence of *S. mansoni* was detected in stool samples. In contrast, the presence of *S. mansoni* was detected in previous studies conducted in other regions of Sudan. It was found to be 2.95%, 5.9%, and 0.9% in Um Asher, White Nile River basin and Al-Gezira, respectively (8, 16, 19). While in Egypt the prevalence was higher (4.3%) (15). These differences because there was no detected *Biomoplaria* snails (the intermediate host of *S. mansoni*) in Al-Fasher due to environmental conditions which may be not suitable for the survival and distribution of *Biomoplaria* snails. In Al-Fasher, especially at the edges of the Foula leak, snails that detected and morphologically identified, according to Sudanese Health Ministry guide of *Schistosomiasis*, were belonged to *Bulinus* snail.

Moreover, hematuria was examined under the microscope and detected in 85.7% of infected patients. Statistically, there was significant correlation between hematuria and presence of *S. haematobium* eggs (P-value = 0.001) which is similar to a study conducted in Al-Lamab Bahar Abiad, Khartoum that showed 77.8% hematuria among infected patients (20). In addition, the prevalence rate of *S. haematobium* according to the water supplies of the study population showed that the infection was most common (11.1%) among those who depend on water delivered by donkey and there was no infection in those who depend on faucets. This might be due to that the water delivered by donkeys may be contaminated with the infective stage of the parasite because, it is untreated water compared with water from faucet which it is treated water. A previous study conducted in North Darfur state showed that the water pollution in the study area is not due to the water sources, but related to the ways used to transport water, stored, and handling (21). In addition, in the present study, there was insignificant association between the infection and age group of the study population which agreed with a previous study (16) and disagreed with a study conducted in Khartoum (20). This variation may be due to differences in sample size, characteristics of the study population and method used in the diagnosis. Also, the present study showed that the children who live in Tambasae and Makraka areas were the most infected, (14.3% and 13.6%, respectively), compared with those who live in Nifasha and Zamzam camps. This could be because hygienic conditions and sanitation systems in the camps were better as the water regularly checked and disposal of human excreta and sewage adequately treated (21). Therefore, providing clean water supplies and adequate sanitary systems are required beside snail control to prevent the spread schistosomiasis in western region of Sudan, esp. AL-Fashir.

### Conclusion

In this study, S. *haematobium* was detected in 6.1% of the urine samples, while no *S. mansoni* was found in the stool samples of the school children at Al-Fasher. In addition, the low prevalence rate of *S. haematobium* was observed in populations who depend on faucets as water sources and live in Nifasha and Zamzam camps. This indicate that the clean water sources and adequate sanitary systems beside snail control are important measurements to control *Schistosomiasis*.

## 4. Limitation

The limitations of this study include the small sample size and the gender of the study population was male. Therefore, further studies with large sample size and collected from both sexes are recommended. To the best of our knowledge, this is the first study that addressed *schistosomiasis* infection in Al-Fasher, capital city of North Darfur.

## Data Availability

All data produced in the present work are contained in the manuscript

